# The impact of COVID-19 in diabetic kidney disease and chronic kidney disease: A population-based study

**DOI:** 10.1101/2020.09.12.20193235

**Authors:** Juan A. Leon-Abarca, Roha S. Memon, Bahar Rehan, Maimoona Iftikhar, Antara Chatterjee

**Author notes:** Medical Doctor, Anaesthesiology and critical care specialist (Antara Chatterjee). **Corresponding author full contact details:** Name: Juan Alonso Leon-Abarca Address: Av. Honorio Delgado 262, San Martín de Porres (Instituto de Investigaciones de la altura). Post code: 15102 City: Lima Country: Peru.

## Abstract

**Background:** The spectrum of pre-existing renal disease is known as a risk factor for severe COVID-19 outcomes. However, little is known about the impact of COVID-19 on patients with diabetic nephropathy in comparison to patients with chronic kidney disease.

**Methods:** We used the Mexican Open Registry of COVID-19 patients 11 to analyze anonymized records of those who had symptoms related to COVID-19 to analyze the rates of SARS-CoV-2 infection, development of COVID-19 pneumonia, admission, intubation, Intensive Care Unit admission and mortality. Robust Poisson regression was used to relate sex and age to each of the six outcomes and find adjusted prevalences and adjusted prevalence ratios. Also, binomial regression models were performed for those outcomes that had significant results to generate probability plots to perform a fine analysis of the results obtained along age as a continuous variable.

**Results:** The adjusted prevalence analysis revealed that that there was a a 87.9% excess probability of developing COVID-19 pneumonia in patients with diabetic nephropathy, a 5% excess probability of being admitted, a 101.7% excess probability of intubation and a 20.8% excess probability of a fatal outcome due to COVID-19 pneumonia in comparison to CKD patients (p< 0.01).

**Conclusions:** Patients with diabetic nephropathy had nearly a twofold rate of COVID-19 pneumonia, a higher probability of admission, a twofold probability of intubation and a higher chance of death once admitted compared to patients with chronic kidney disease alone. Also, both diseases had higher COVID-19 pneumonia rates, intubation rates and case-fatality rates compared to the overall population.

## Introduction

The prevalence of SARS-CoV-2 and the disease it causes (COVID-19) is increasing worldwide. With more than 17 million cases worldwide, COVID-19 is a pandemic declared by the World Health Organization (WHO) with the first recorded cases In Wuhan, China. Since then, it has spread rapidly to other areas of the world. People who are infected with the virus may develop complications that lead to pneumonia, intubation, ICU admission, and even death. The WHO states that elderly and people who are already suffering from pre-existing medical conditions such as chronic kidney disease (CKD), diabetes, or cardiovascular disease are at a higher risk to develop a severe disease from their exposure to COVID-19 with an increased rate of morbidity and mortality **[1-2]**.

The way by which the SARS-CoV-2 virus is believed to bind to the host target cell is similar to the previous coronavirus (SARS-CoV) which binds to the target cell via angiotensin-converting enzyme 2 (ACE2). ACE2 is expressed by the epithelial lining of the lungs, blood vessels, and kidneys. The expression of ACE2 is increased in patients suffering from CKD and hence the main treatment for both these diseases is through ACE inhibitors which further increases the expression of ACE2 **[3]**. CKD patients often suffer from multiple comorbidities, including diabetes. Patients with type 1 and type 2 diabetes have a substantially increased expression of ACE2 as they are treated with ACE inhibitors and angiotensin II type-I receptor blockers (ARBs), thus infection with SARS-CoV-2 would be facilitated by the increased expression of ACE2 in CKD and their treatment with ACE2 inhibiting drugs might increase the risk and fatality of developing severe COVID-19 **[4]**.

Several studies have also stated that the presence of comorbidities has been associated with a 3 to 4 fold increase in the risk of developing acute respiratory distress syndrome leading to intubation and/or ICU admission and/or development of pneumonia in patients with SARS – CoV **[5]** and Middle East Respiratory Syndrome coronavirus (MERS-CoV**)[6]** both of which belong to the same family of the beta-coronavirus genus **[7]**. These previous coronavirus infections, SARS-CoV and (MERS-Co-V), have infected more than 10,000 people in the past 2 decades, with mortality rates of 10% and 37% respectively **[8-9]**. COVID-19, being far more contagious than these illnesses combined has a greater predisposition to respiratory failure, intubation, COVID-19 pneumonia, ICU admission or even death in highly susceptible patients due to its highly infectious nature and rapid spread **[10]**. Hence an updated analysis with a significant sample size is urgently warranted as there is scarce data of the impact of COVID-19 on CKD patients. The present analysis will explore if there are differences in COVID-19 outcomes in patients with CKD with any cause other than diabetes and those patients with diabetic nephropathy. The results might aid in renal patient management and help in the development of policies for prevention and response to COVID-19 and its critical outcomes.

## Methods

### Data source

We used the Mexican Open Registry of COVID-19 patients 11 to analyze anonymized records of those who had symptoms related to COVID-19 and consulted a healthcare centers either in two broad conditions: 1) Acute respiratory distress symptoms or severe respiratory infections 2) Mild respiratory infections that could be treated in the outpatient clinic. A nasopharyngeal RT-PCR assay for SARS-CoV-2 was taken for every patient with severe features and only one every out of ten patients with mild cases were sampled. According to Mexican COVID-19 guidelines, every patient who had symptoms related to COVID-19 in the past 7 days is considered under suspicion of the diseases until proven otherwise.

### Data analysis plan

In this analytical cross-sectional study we use the individual records of adult Mexican people who have a registry of defined RT-PCR (either positive or negative), sex, age in years and both place of residence and place at birth. Also, we included patients who had a record of chronic kidney disease. Other diseases that might act as severe COVID-19 risk factors such as hypertension and asthma were excluded to accurately represent the impact of diabetes in patients living with CKD. After these inclusion and exclusion criteria, 2092 patients with CKD only and 742 patients with diabetic nephropathy (defined as the patient who had diabetes and chronic kidney disease) were included. We used robust Poisson regression models to relate sex and age to each of the six outcomes: SARS-CoV-2 infection, development of COVID-19 pneumonia, hospitalization, intubation, Intensive Care Unit admission and death. These regressions were used to estimate adjusted prevalences (aP) and adjusted prevalence ratios (aPR), which were compared through Bonferroni corrections. Binomial regression models were performed for those outcomes that had significant results, considering the same independent variables and their interactions. Interactions included sex and diabetes, sex and CKD, age and diabetes, and age and CKD. Predictive marginal analysis was performed to plot the probabilities along 95% confidence bands to analyze the change in probability along age for both chronic kidney disease and diabetic nephropathy. Only to compare the probability of achieving each of the six COVID-19 outcomes for both diseases against the total population, we used 438096 cases consisting of the entire dataset without both diseases to estimate a baseline probability. The data was analyzed through the STATA 14 program considering a p value of 0.05 as the statistically significant threshold.

### Data management statement

Processed data files used in the present study are available at request. The authors follow the STrengthening the Reporting of OBservational studies in Epidemiology (STROBE) statement for data collection, analysis and reporting of results **[12]**.

## Results

Summary characteristics of the study population are presented in **Table 1**. The mean age of the population was 44.4 years (95% CI: 43.74-45.15) and patients with diabetic nephropathy were in average 13.8 years older than CKD patients. The male to female ratio was 1.2527 and there were no significant differences in sex proportion between the two examined pathologies (p>0.05). The observed prevalence of chronic kidney disease in the general population was 0.51% (95% CI: 0.48-0.53%) while the prevalence of diabetic nephropathy was 2.69% (95% CI: 2.51-2.89%). Patients with diabetic kidney disease were 10.7% more likely to be infected with SARS-CoV-2 compared to CKD patients, doubled their rate of COVID-19 pneumonia (107.9% more), doubled their rate of intubation (103.4% more), had a 52.6% higher ICU admission rate and had a 27.14% higher case-fatality rate (p< 0.05 for each crude prevalence comparison). There were no differences between diseases in time from symptom onset to healthcare consult and time from admission to death, although a tendency of a shorter span in days was seen amongst patients with diabetic nephropathy (p>0.05).

**Table 1.**
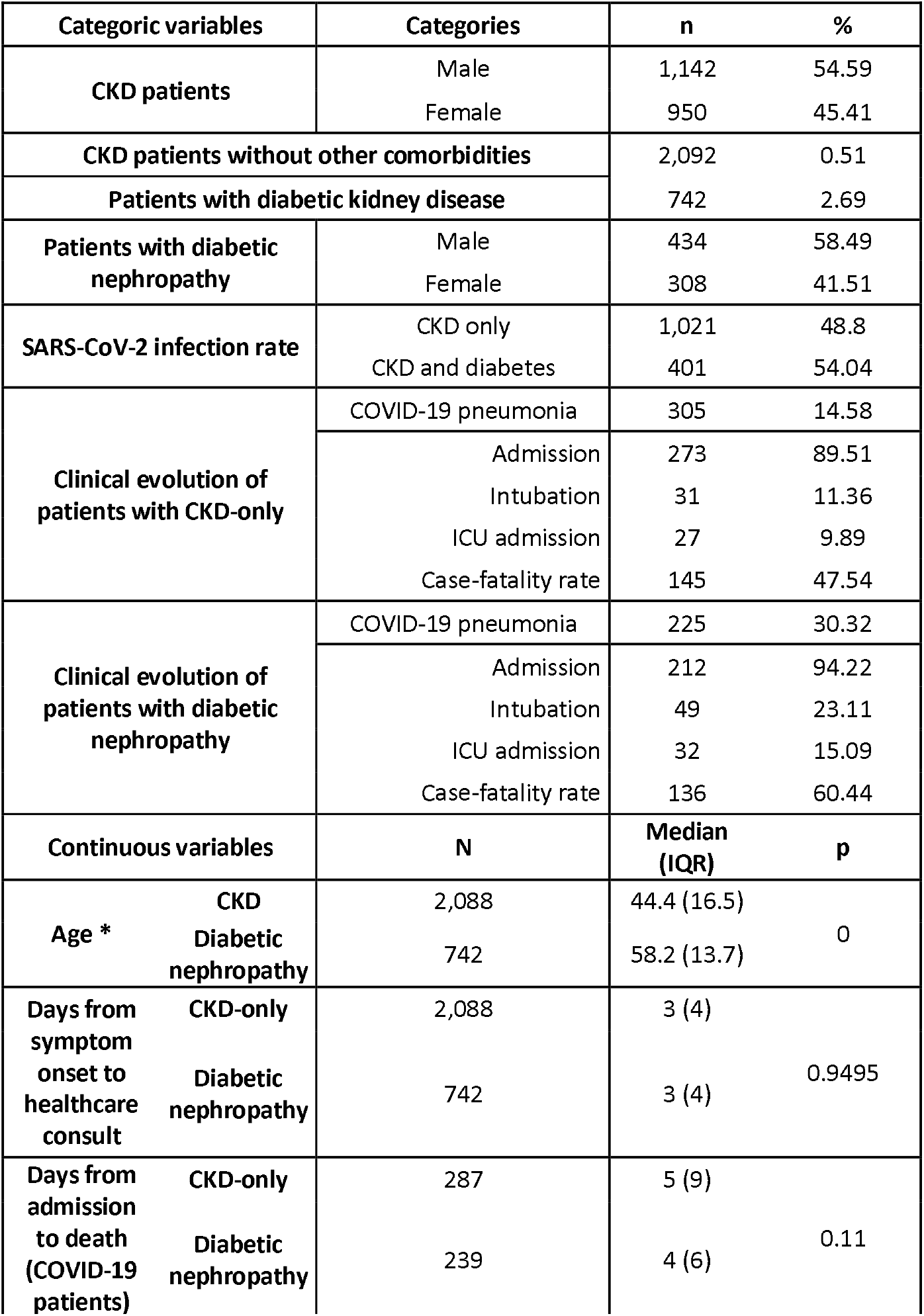
Summary characteristics of the study population. * Mean and Standard deviation instead of Median and Interquartile Range (IQR). Total population to obtain prevalence rates n = 413,372. CKD: chronic kidney disease. DBM: Diabetes

The adjusted prevalence ratios **(Table 2)** revealed that those with diabetic nephropathy had no difference in SARS-CoV-2 infection compared to patients with CKD-only but had a 87.9% excess probability of developing COVID-19 pneumonia, a 5% excess probability of being admitted, a 101.7% excess probability of intubation and a 20.8% excess probability of a fatal outcome all due to COVID-19 pneumonia (p< 0.01 for all cases). Of note, the adjusted prevalence of admission due to COVID-19 pneumonia was high in both CKD-only patients and patients with diabetic nephropathy.

**Table 2.** Adjusted prevalence ratios (aPR) and adjusted prevalences (aP) by age and sex for patients with and without diabetes shown along 95% CI’s. Highlighted: statistically significant results at p< 0.05

| Outcomes | Diabetic nephropathy<br>versus CKD alone |  | CKD + DBM |  | CKD only |  | n |
| --- | --- | --- | --- | --- | --- | --- | --- |
|  | aPR | 95% CI | aP | 95% CI | aP | 95% CI |  |
| SARS-CoV-2<br>infection | 1.068 | 0.98-1.16 | 50% | 47.6-52.0 | 53.18% | 49.4-56.9 | 2,830 |
| COVID-19<br>pneumonia | 1.879 | 1.58-2.17 | 30.15% | 26.6-33.7 | 16.05% | 14.4-17.7 | 2,830 |
| Admission | 1.056 | 1.001-1.11 | 95.54% | 92.2-98.9 | 90.50% | 87.1-93.9 | 530 |
| Intubation | 2.017 | 1.18-2.85 | 23.10% | 17.4-28.8 | 11.45% | 7.7-15.2 | 485 |
| ICU admission | 1.653 | 0.81-2.49 | 15.71% | 10.5-20.9 | 9.50% | 6.0-12.9 | 485 |
| Fatalities | 1.208 | 1.01-1.40 | 59.88% | 53.3-66.5 | 49.55% | 43.8-55.3 | 530 |

A more detailed analysis through the age continuum up to 90 years using probability trends is presented in **Image 1**. Patients with diabetic nephropathy and CKD had a higher probability of developing COVID-19 pneumonia once infected with SARS-CoV-2, however this association only was present for patients up to 70 years (in the case of CKD) and 76 years (in the case of diabetic nephropathy (p< 0.05) (**Image 1 – Panel A)**. The probability of admission was statistically significantly lower in patients without both diseases up to 60 years old in the case of CKD and 65 years in the case of patients with diabetic nephropathy **(Image 1 – Panel B)**. The intubation rate was higher only for patients with diabetic nephropathy compared to the general population until 76 years old, however there was no statistically significant differences (p>0.05) in intubation rates between patients with CKD and the general population. Also, patients with diabetic nephropathy had higher intubation rates (p< 0.05) compared to CKD-only patients between 40 to 70 years old (**Image 1 –Panel C)**. The case-fatality-rate for patients with COVID-19 pneumonia was always higher for both diseases up until 81 years old (**Image 1 – Panel D)**.

**Image 1.**
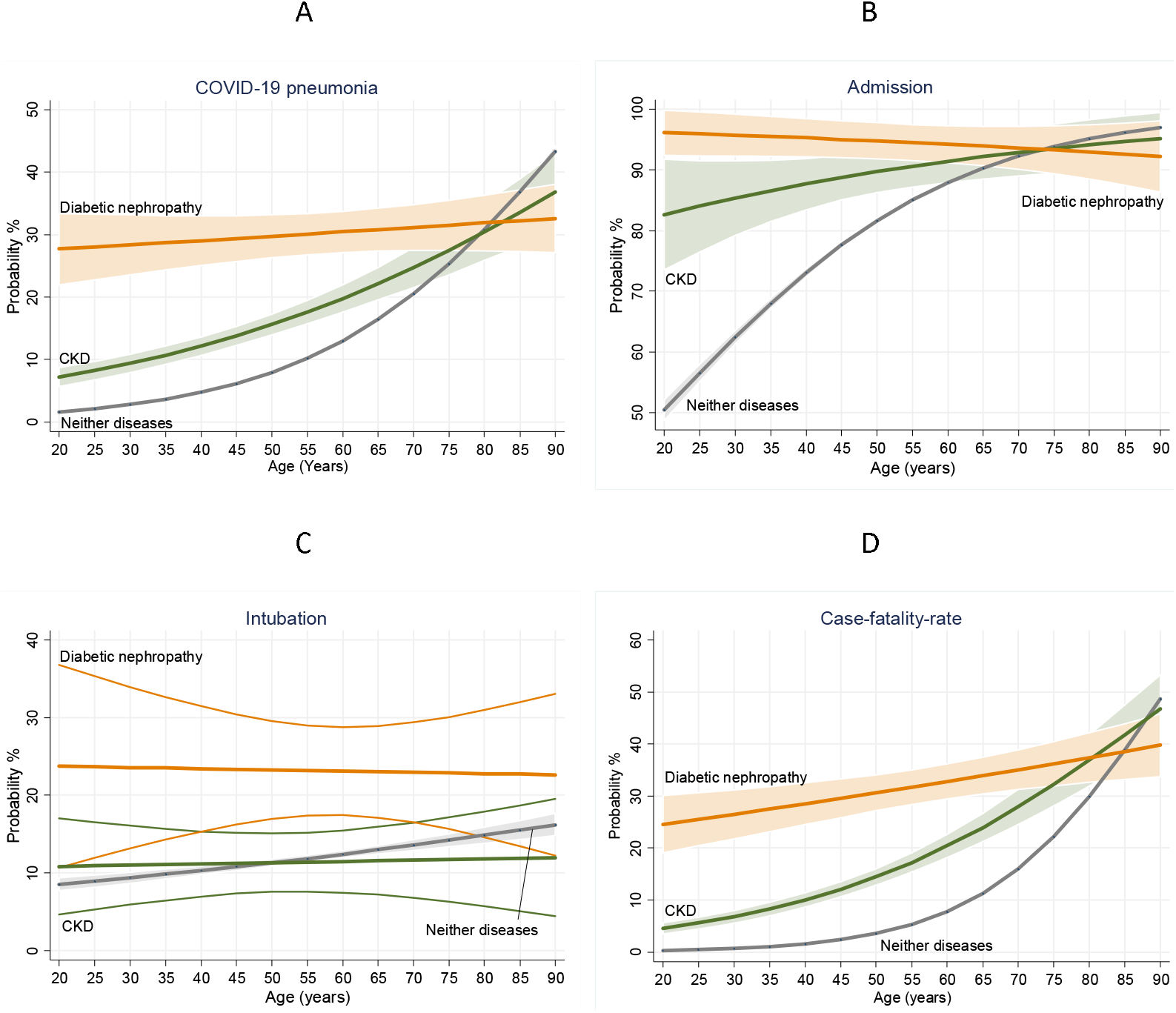
Probability trends with 95% confidence bands along age for COVID-19 outcomes.

## Discussion

In our retrospective cross-sectional study of Mexican adults, the principal finding was that patients with diabetic nephropathy had nearly a twofold rate of COVID-19 pneumonia, a higher probability of admission, a twofold probability of intubation and a higher case-fatality rate once admitted compared to patients with chronic kidney disease alone. Also, both diseases had higher COVID-19 pneumonia rates, intubation rates and case-fatality rates compared to the overall population.

Our analysis revealed that almost one out of six cases with isolated CKD had COVID-19 infection. In a previous study, nearly half of the patients (47.6%) diagnosed with COVID-19 had underlying CKD **[13]**. In addition, the viral illness took a severe course of progression in CKD patients in our study. Similar findings have been reported in the current literature. The information on the Centers for Disease Control and Prevention (CDC), with recent updates as of 25^th^ June 2020, reports a higher risk of severe COVID-19 infection in CKD cases, regardless of the stage of kidney disease **[14]**. In a metaanalysis conducted on the preliminary data, CKD was concluded to be a significant predictor of the clinical outcomes associated with COVID-19 infection albeit individual studies included in this metaanalysis reported no such association **[15]**.

Furthermore, we assessed and compared the severity of clinical course of COVID-19 in cases with diabetic nephropathy to that in CKD cases without diabetes by evaluating various factors. The rate of contracting COVID-19 infection was 87.9% higher in diabetic nephropathy versus CKD patients. Our analysis also highlighted the more severe clinical course from COVID-19 in patients with diabetic nephropathy. They had nearly a twofold rate of COVID-19 pneumonia, a higher probability of admission, a twofold probability of intubation and a higher chance of death once admitted, compared to patients with CKD alone. Consistently, the CDC has raised concerns on the increased risk of serious illness with diabetes mellitus (DM) type 1 and gestational DM **[14]**. A recent study on the impact of comorbidities on the disease course and outcomes of COVID-19 concluded greater disease severity in contrast to those without these comorbid conditions. Amongst all, DM was found as the second most prevalent pre-existing condition in the patient population and posed significant risk in achieving ICU admission, invasive ventilation and death. Furthermore, this study also focused on the frequent co-existence of the comorbidities which added individually and escalated the risk of poorer outcomes **[16]**. Since CKD and DM frequently coexist as diabetic kidney disease (DKD), it is plausible to expect worse outcomes in terms of vulnerability to serious viral illnesses **[17]**. Our study provides robust support to the findings of these studies as more than half of the of population with diabetic nephropathy (54.04%) had COVID-19 infection as compared to 48.8% infection rate in patients with just CKD and furthermore, a third of patients with diabetic nephropathy infected with SARS-CoV-2 developed COVID-19 pneumonia, with nearly all of them needing hospitalization (94.22%).

DM and CKD are common diseases that exhibit synergistic associations with premature mortality. CKD has a significant worldwide prevalence affecting 7.2% of the global adult population with the number dramatically increasing in the elderly **[18]**. Although the causes are various, diabetes is the most common cause of CKD in the United States and an increasing cause of the same worldwide. Diseases of the kidney are a common finding in people with DM, with up to half demonstrating signs of kidney damage in their lifetime **[19-20]**. A variety of forms of kidney disease can be seen in people with diabetes, including diabetic nephropathy, ischemic damage related to vascular disease and hypertension, as well as other renal diseases that are unrelated to diabetes. The pathogenesis is multifactorial involving adaptive hyperfiltration, advanced glycosylated end-product synthesis (AGES), nephrin expression and impaired podocyte-specific insulin signaling. Treatments focus on lifestyle interventions including control of hyperglycemia, hypertension and hyperlipidemia as well treatment of complications and preparation for renal replacement therapy **[18]**. Kidney disease can be a particularly devastating complication, as it is associated with significant reductions in both length and quality of life **[21]**. Diabetic patients usually have a significant reduction of the forced vital capacity and forced expiratory volume during the first second **[22]**. These lung changes could predispose diabetic patients with COVID-19 to have a worse prognosis. In line, it has been reported that elevated levels of glycosylated hemoglobin (HbA1c) is associated with an increase of 60% in risk of hospitalization and severity of pneumonia in bacterial infections **[23]**. Hyperglycemia is considered an independent factor which increases the risk of death and worsens the prognosis **[24-25]**. Poor glucose control in many diabetics could promote glycosylation of angiotensin-converting enzyme 2 (ACE 2), the gateway for SARS-CoV-2 in the host **[26]**.

Plausible explanations could be provided for a greater incidence and severity of COVID-19 infection in CKD or diabetic nephropathy cases. A simpler yet a practical one could be that these patients with the need to undergo routine dialysis are at a risk of coming in at least indirect contact with infected cases, contracting the infection. However, CKD could also up the chances of infection pathophysiologically. It is well-known that kidney disease especially diabetic nephropathy **[27]**, is associated with a proinflammatory state and functional deficits in both the innate and adaptive immune systems **[28]**. Furthermore, immune-modulating therapies used to treat it can add to the immune dysregulation **[29]**. In addition, angiotensin converting enzyme 2 (ACE2) receptors are overexpressed in renal tubular cells of CKD patients **[30]**. Since the entry of SARS-CoV-2 in the kidney cells is mediated by ACE2 receptors, the overexpression of these receptors together with the underlying immune dysfunction could leave the individual vulnerable to COVID-19 infection and its fatal outcomes. Together with increased incidence, the dysfunction of immune system could also be speculated as the mediator of worsening of clinical outcomes **[29-31]**.

Regardless of COVID-19, CKD has been reported to precipitate outpatient pneumonia **[32]** as well as a more severe one requiring inpatient treatment **[33]**. One study suggests immunosuppression and a higher co-existence of comorbid conditions (diabetes for example), the root cause of the higher incidence of COVID-19 pneumonia in CKD cases on hemodialysis **[34]**. From symptom onset, both patient populations in our study took a median of 3 days before consulting healthcare, however higher rates of ICU admission and subsequent intubation were seen in patients with diabetic nephropathy. The disease was seen to be rapidly progressive in cases with diabetic nephropathy as they survived a median of 4 days following admission compared to 5 days in isolated CKD cases, although no significant differences were found.

Finally, a high mortality was seen in all CKD patients, albeit the rate was higher in diabetic nephropathy cases. In one study conducted on COVID-19 patients in a Washington hospital, nearly half of them with CKD, higher mortality was reported amongst them **[13]**. Mortality rates in CKD patients with COVID-19 were also studied in tertiary hospitals in Wuhan, China **[35]**. Following adjustment for baseline characteristics like age and sex, abnormal renal functions as predicted by elevated serum creatinine and urea along with proteinuria and hematuria and acquired acute kidney injury became strong independent risk factors for in-hospital death. Considering a higher prevalence and co-existence of these risk factors in each of CKD and diabetic nephropathy cases, the higher mortality seen in our study is reasonable **[35]**. In one study, higher morbidity and mortality was reported in diabetic kidney disease cases. The study attributed it to the complications developed in the course of COVID-19 infection as a result of generalized immunosuppression secondary to chronic systemic inflammation seen in this cohort of patients **[27]**.

The aim of our study was to study in detail both CKD and diabetic nephropathy as those are two common diseases amongst patients with chronic renal pathologies. We found that diabetes does have a significant additional impact on the rate of infection, intubation, ICU admissions and case fatality rate due to COVID 19 compared to patients with CKD alone. We can then confirm the hypothesis that the morbidity and mortality of patients infected with SARS-CoV-2 is significantly higher in patients with diabetic kidney disease than those with CKD alone, possibly due to the added effects of chronic inflammation and immune dysfunction. The rates of SARS-CoV-2 infection, ICU admissions, case-fatality rates are higher in diabetic kidney disease than in CKD patients alone, while rates of pneumonia and intubation double.

Further speculation could be drawn from our results, as patients with diabetic nephropathy and CKD have a higher probability of developing COVID-19 pneumonia once infected with SARS-CoV-2, but this association only holds for patients up to 70 years (in the case of CKD) and 76 years (in the case of diabetic nephropathy). The intubation rate seems higher only for patients with diabetic nephropathy compared to the general population until 76 years old, however there does not seem to be a statistically significant difference (p>0.05) in intubation rates between patients with CKD and the general population. The case-fatality-rate is always higher for both diseases up until 81 years old. Further research is needed to establish why COVID pneumonia development was more probable till the age 76 and not thereafter. Further research regarding this relationship and its clinical management is warranted.

Few limitations should be considered. In view of the publicly available data, we could not have access to patients’ clinical records including laboratory test reports. Thus, the severity of the underlying kidney disease could not be taken into consideration in the study. Due to similar reasons, it was unclear if patients received treatment for diabetes or were under any kind of renal replacement therapy. Secondly, given the observational nature of our study, the associations found may not be causal. Although we were able to adjust for multiple confounding factors, we cannot rule out unmeasured or residual confounding. Furthermore, a limited number of studies have assessed the clinical picture of COVID-19 in patients with diabetic nephropathy, conducted in a few countries only. Thus, a detailed comparison and contrast could not be drawn between the findings of our study and those previously conducted. Other than a few hypotheses on the poorer COVID-19 endpoints, our study did not aim to portray a robust clinical picture of COVID-19 outcomes in cases with multiple comorbid conditions.

Our manuscript calls for further research in this area as we conclude alarming findings. The greater severity of the COVID-19 disease course in CKD and diabetic nephropathy patients is then confirmed albeit the pure clinical and laboratorial picture remains unclear. Future research in other settings could help fill these gaps in the knowledge and would help formulate guidelines of care in these complicated cases. In addition, future research should incorporate population with multiple underlying comorbidities so that a holistic well-powered assessment of the effects of mounting comorbidities on the COVID-19 outcomes could be evaluated.

## Data Availability

Processed data files used in the present study are available at request. The authors follow the STrengthening the Reporting of OBservational studies in Epidemiology (STROBE) statement for data collection, analysis and reporting of results.

